# Longitudinal Symptom Burden and Pharmacologic Management of Catatonia in Autism with and without Profound Impairment: An Observational Study

**DOI:** 10.1101/2024.09.05.24312724

**Authors:** Joshua Ryan Smith, Seri Lim, Snehal Bindra, Sarah Marler, Bavani Rajah, Zachary J. Williams, Isaac Baldwin, Nausheen Hossain, Jo Ellen Wilson, D. Catherine Fuchs, James Luccarelli

## Abstract

**Introduction:** Catatonia is a highly morbid psychomotor and affective disorder which can affect autistic individuals with and without profound impairment. Catatonic symptoms are treatable with pharmacotherapy and electroconvulsive therapy, but the longitudinal effectiveness of these treatments has not been described.

**Methods:** We conducted a prospective observational cohort study of patients with autism and co-morbid catatonia who received outpatient care in a specialized outpatient clinic from July 1^st^, 2021 to May 31^st^, 2024. Data investigating pharmacologic interventions, and clinical measures including the Bush Francis Catatonia Rating Scale (BFCRS), Kanner Catatonia Severity Scale (KCS), Kanner Catatonia Examination (KCE), and Clinical Global Impression – Improvement (CGI-I) were collected.

**Results:** Forty-five patients were identified with 39 (86.7%) meeting criteria for profound autism. All patients received pharmacotherapy. 44 (97.8%) were treated with benzodiazepines with a mean maximal daily dose of 17.4 mg (SD=15.8) lorazepam equivalents. Thirty-five patients (77.8%) required more than one medication class for treatment. Fourteen patients (31.1%) attempted to taper off benzodiazepines during the study period; of these, 5 patients (11.1%) were successfully tapered off, and the remaining 9 (17.8%) discontinued the taper due to a return of catatonic symptoms. Statistically significant improvement was observed across all clinical domains except the KCS. However, the majority remained symptomatic over the study period.

**Conclusions:** Despite clinical improvements while receiving the gold standard for psychopharmacologic management of catatonia, chronic symptoms remained for the majority of catatonia patients over the study period, and few were able to taper and discontinue benzodiazepine treatment.

**Lay Summary:** Catatonia is a condition which can significantly impact the quality of life for autistic individuals with and without profound impairment. Symptoms of catatonia such as aggression and self-injury can improve with treatment. However, over the three-year period of this study, most patients experienced residual symptoms, and few could discontinue psychotropic medication.

## Introduction

Autism is a heterogeneous neurodevelopmental condition characterized by difficulties in social interaction and communication along with restricted/repetitive patterns of behaviors and interests.^1^ Autism affects 1-2% of the population worldwide. A subset of autistic individuals are classified under the newly-proposed category of “profound autism”,^2^ a developmentally-stable phenotype in which individuals older than 8 years of age require 24-hour access to an adult who can care for them, are unable to live completely alone in a residence, and are not able to take care of basic or adaptive needs.* Operational diagnostic criteria for profound autism have been proposed in which an autistic individual meets criteria for (a) moderate-to-profound intellectual disability and/or (b) “nonverbal” or “minimally verbal” status^2^ due to cognitive or language impairment (i.e., excluding pure motor speech impairment).^5^ Overall, it has been estimated that approximately 26% of autistic individuals in the United States meet these operational criteria for profound autism.^6^

Individuals with profound autism are at an elevated risk for co-occurring psychiatric conditions, caregiver distress, aggression, residential placement, and hospitalization.^7–9^ One co-occurring psychiatric condition which often presents with aggression and may occur at a higher frequency in profound autism populations is catatonia.^10,11^ Catatonia is a psychomotor syndrome with affective domains and distinct physical examination findings, which can affect individuals across the lifespan.^12–14^ A recent meta-analysis by Vaquerizo-Serrano and colleagues found that 20.2% of autistic individuals had features of catatonia, with 10.4% meeting full criteria for catatonia. The most common symptoms of catatonia in autism were new-onset speech impairment, negativism, hyperactivity, and aggression deviating from the individual’s baseline.^10^ Moreover, recent literature has suggested that recurrent self-injury is a symptom along the catatonia spectrum for autistic individuals with profound impairment.^15^ While classically described features of catatonia (such as staring, posturing, and waxy flexibility) do occur in autism, externalizing (for example: aggression, impulsivity, self-injury) and hyperactive symptoms are more common. Thus, there is an increased risk of a missed diagnosis given the phenotypic variability between catatonia occurring in neurotypical individuals and those with autism.

Use of electroconvulsive therapy (ECT) and benzodiazepines has demonstrated efficacy in the treatment of catatonia for both neurodivergent and neurotypical adults and children.^16–18^ While some inpatient data is available,^19^ there is a need for longitudinal outpatient studies of catatonia in autism to define the course of the disorder.^20^ In addition, there is limited data available to guide the use of other pharmacologic agents beyond benzodiazepines in the population of autistic individuals with catatonia, particularly in the outpatient setting.^21,22^ Thus, we conducted a prospective observational longitudinal study of autistic individuals, with and without profound impairment, who received care in a specialized neurodevelopmental catatonia clinic. The aim of our study was to provide longitudinal diagnostic, pharmacologic, and therapeutic data to assist clinicians in providing care for this high need population.

## Methods

### Study Population

Utilizing *Strengthening the Reporting of Observational Studies in Epidemiology* (STROBE) guidelines,^23^ we conducted an observational cohort study of patients with autism and co-morbid catatonia who received outpatient care in a specialized clinic at Vanderbilt University Medical Center. Patient information was collected longitudinally from July 1^st^, 2021 to May 31^st^, 2024. Patients were included if they were seen at least twice in the outpatient clinic and were diagnosed with both autism and catatonia. The diagnosis of autism was confirmed by the presence of a previous community diagnosis and/or review of historical medical records. The diagnosis of profound autism was confirmed during longitudinal clinical care, following the definitions as described. This study was reviewed by the Vanderbilt University IRB (#170317) with a waiver of informed consent from participants. This study is a continuation of our previous work investigating the use of ECT for adults and children with autism and intellectual disability,^17,18,24^ and pediatric catatonia more broadly.^11,13,21^ Thus, data from some patients in this cohort has been previously reported in terms of diagnostic categorization and therapeutic applications in catatonia. However, the specific longitudinal clinic data presented here is novel and has not yet been reported.

### Measures

Therapeutic data including psychopharmacologic interventions, use of ECT, and catatonia assessment measures (the Bush Francis Catatonia Rating Scale [BFCRS],^25^ Kanner Catatonia Severity [KCS], and Kanner Catatonia Examination [KCE]),^26^ were recorded over the course of the study period. The BFCRS,^27^ the most cited paper in the field of catatonia,^28^ consists of a 14-item Bush Francis Catatonia Screening Item (BFCSI) and a full 23-item BFCRS which measure a greater number of signs. The Kanner Catatonia Rating Scale includes the KCS severity assessment and the KCE, a standardized examination.^26^ To determine if clinical improvement occurred over the course of longitudinal clinical care, total BFCRS scores were collected at the time of clinic intake (patients presenting for clinical care and receiving a diagnosis of catatonia upon intake) or, when the diagnosis of catatonia was first given (for patients receiving a diagnosis of catatonia while already followed in-clinic). Secondarily, KCS and KCE scores were collected in the same manner, The clinical global impression – improvement (CGI-I) scale^29^ was determined based on the patient’s improvement since clinic intake and was calculated by the primary treating psychiatrist (author JRS).To investigate the impact of catatonia on families and autistic individuals of varying socioeconomic backgrounds, data regarding healthcare payers (Medicaid, private insurance, self-pay) were also collected.

### Data Collection

Demographics, co-occurring medical and psychiatric conditions, the number of failed psychiatric medications, frequency and severity of catatonic symptoms as measured by catatonic assessment measures (BFCRS, KCE, KCS), use of ECT, mortality, and pharmacologic initiation and/or tapering were recorded prospectively during clinic visits. Catatonic signs and symptoms assessed at baseline (particularly those such as verbigeration and stereotypy, which could overlap with features of autism) were verified with patients’ caregivers as representing a noticeable difference from the patient’s typical behavior before the presumed onset of catatonic symptoms.

Given the heterogeneity of benzodiazepines used in the treatment of catatonia in this population, dosing of benzodiazepines was converted into lorazepam equivalents, with 0.5 of clonazepam, 5 mg of diazepam, 10 mg of clobazam considered equivalent to 1 mg of lorazepam.^30,31^ Length of outpatient follow up was calculated from the date of clinic intake until either clinic discharge, the end of the study period, or death.

To specifically address the prevalence of abnormal physical examination findings, the frequency of catatonic symptoms as measured by the BFCRS, KCS, and KCE was also collected. The specific clinical signs measured by each scale are listed in Supplemental tables 1 and 2. For the KCS specifically, positive items (scoring a 2 or more on the KCS, and a score of 1 on the KCE) were included when determining symptom or physical examination finding frequency. Notably, self-injury on the KCS is reported as mannerism or stereotypy with accompanying self-injury (indicated by a score of 4 or higher on each item). Thus, symptom severity was also reported in these two domains to best capture the presence of self-injury.

The KCS and KCE report catatonic symptoms such as self-injury and incontinence which are not reported in the BFCRS. The BFCRS does not separately assess for reduced food and fluid intake but combines these two measures. The BFCRS also does not assess for magnetism or the metronome test on physical examination, both of which are present on the KCE. For ease of administration for children and neurodivergent populations, the KCE combines automatic obedience and ambitendency into one physical examination finding referred to as command-verbal. Notably, affective symptoms of catatonia are not captured by either measure.

### Statistical Analyses

Statistical analyses were performed using Python (Version 3.12.4; Python Software Foundation) and R (Version 4.4.1; R Core Team, 2024). The primary statistical endpoint was the change in BFCRS total score between time of first diagnosis and time of final clinic appointment. This endpoint was chosen to permit comparison with prior research.^32,33^

To determine if improvement in BFCRS, KCS, and KCE scores occurred over the study period, the most recent BFCRS, KCS, and KCE scores were collected from the last documented clinical note of each participant within the study period. Hierarchical models were then estimated to approximate the linear trends in catatonia symptom improvement between baseline clinical assessment and the end of the follow-up period. All available BFCRS, KCS, and KCE data from either the baseline or most recent timepoints were modeled using outcome-specific intercept-only hierarchical linear or ordered-probit models, with one model constructed for each catatonia outcome measure. Hierarchical linear models (used to model BFCRS and KCS scores) were fit using restricted maximum likelihood estimation in the *lme4* R package,^34^ whereas the single hierarchical ordered-probit model^35^ was fit using a Laplace approximation to maximum likelihood, as implemented in the *clmm* function in the *ordinal* R package,^36^ and both model types utilized Wald approximations for fixed-effect *p*-values and profile likelihood for the calculation of 95% confidence intervals. Participants with a BFCRS, KCS, or KCE at only one time point still had their data included in these models, as full information maximum likelihood estimation allowed for such participants to contribute to the marginal effects of each model. Effects were quantified as the raw mean difference in BFCRS/KCS score points over the course of the study (denoted using the regression slope, β), as well as the standardized mean difference (*d*/*d_ord_*) defined as the reduction in BFCRS/KCS/KCE outcome standard deviation units (or latent probit units for *d_ord_*) between timepoints 1 and 2. Additionally, CGI-I scores for all participants were compared to a baseline score of 4 (“No change”) using a one-sample ordinal statistical test (Wilcoxon signed-rank test; rank-biserial correlation *r_c_* used as a standardized effect size). A two-tailed *p*-value of <.05 was used to determine statistical significance, and Bonferroni-Holm corrections were performed to control the familywise error rate of the four statistical tests (test of timepoint parameters for BFCRS/KCS/KCE hierarchical models, CGI-I Wilcoxon test).

## Results

### Demographics, autism diagnoses, and length of outpatient follow up

Overall, observational data was collected for 45 patients with autism and catatonia (Table 1). The mean age of patients in the clinic at index visit was 15.6 (*SD*=7.9) years [Mdn=16.0, range 6.0–31.0]. With regard to sex assigned at birth in the 45 patients, 31 (68.9%) were male and 14 (31.1%) were female. Medicaid was the primary payor for 30 (66.7%) patients, while 12 (26.7%) were covered by private insurance, and 3 (6.7%) were self-pay. Profound autism (defined using the Lancet commission operational criteria)^2^ was highly prevalent in this cohort (*n*=39, 86.7%). Additionally, 41 patients (91.1%) met criteria for autism with co-occurring intellectual disability.

**Table 1:**
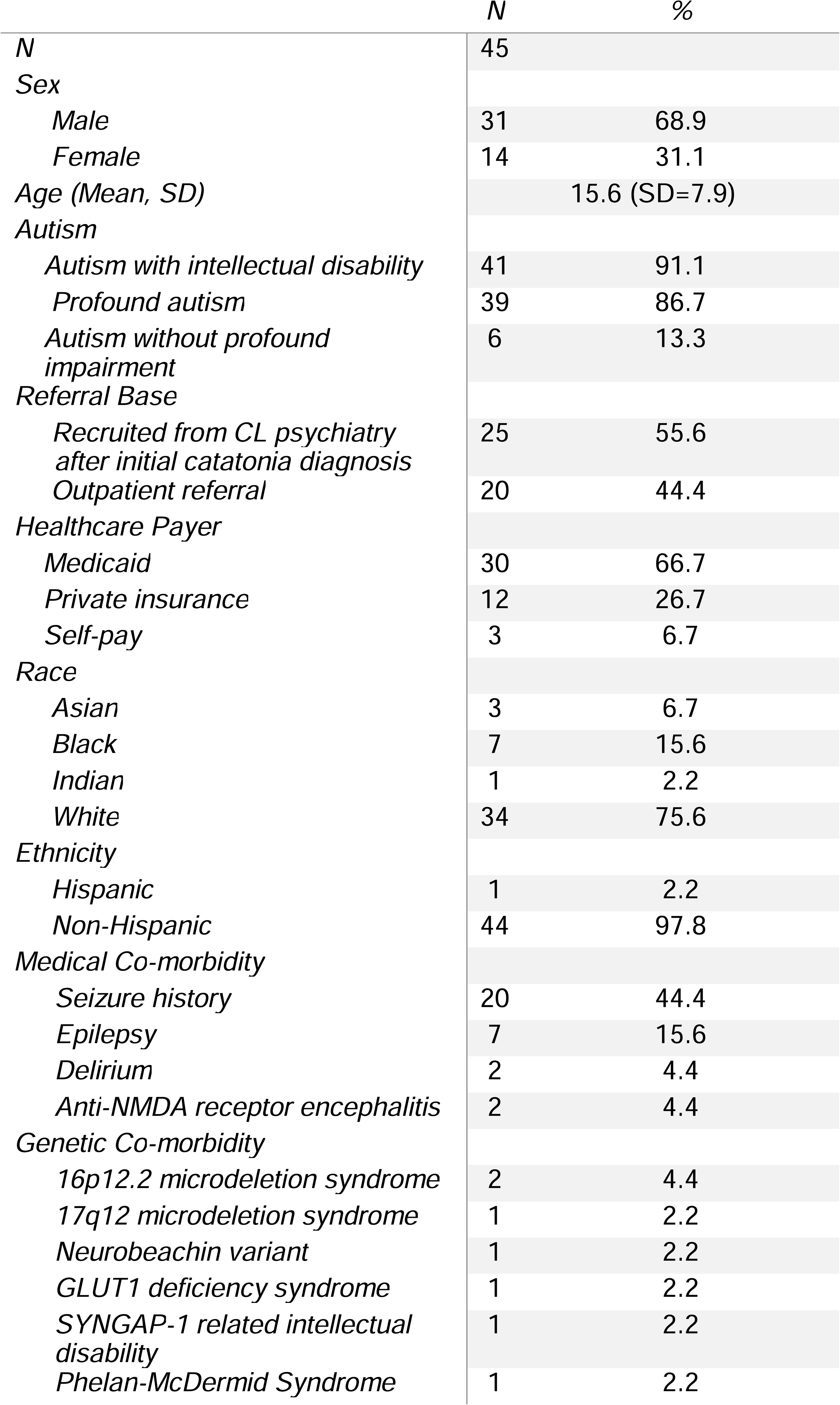
Demographics, genetic and medical co-morbidity for patients with autism and catatonia.

|  | N | % |
| --- | --- | --- |
| *N* | 45 |  |
| *Sex* |  |  |
| *Male* | 31 | 68.9 |
| *Female* | 14 | 31.1 |
| *Age (Mean, SD)* | 15.6 (SD=7.9) |  |
| *Autism* |  |  |
| *Autism with intellectual disability* | 41 | 91.1 |
| *Profound autism* | 39 | 86.7 |
| *Autism without profound impairment* | 6 | 13.3 |
| *Referral Base* |  |  |
| *Recruited from CL psychiatry after initial catatonia diagnosis* | 25 | 55.6 |
| *Outpatient referral* | 20 | 44.4 |
| *Healthcare Payer* |  |  |
| *Medicaid* | 30 | 66.7 |
| *Private insurance* | 12 | 26.7 |
| *Self-pay* | 3 | 6.7 |
| *Race* |  |  |
| *Asian* | 3 | 6.7 |
| *Black* | 7 | 15.6 |
| *Indian* | 1 | 2.2 |
| *White* | 34 | 75.6 |
| *Ethnicity* |  |  |
| *Hispanic* | 1 | 2.2 |
| *Non-Hispanic* | 44 | 97.8 |
| *Medical Co-morbidity* |  |  |
| *Seizure history* | 20 | 44.4 |
| *Epilepsy* | 7 | 15.6 |
| *Delirium* | 2 | 4.4 |
| *Anti-NMDA receptor encephalitis* | 2 | 4.4 |
| *Genetic Co-morbidity* |  |  |
| *16p12.2 microdeletion syndrome* | 2 | 4.4 |
| *17q12 microdeletion syndrome* | 1 | 2.2 |
| *Neurobeachin variant* | 1 | 2.2 |
| *GLUT1 deficiency syndrome* | 1 | 2.2 |
| *SYNGAP-1 related intellectual disability* | 1 | 2.2 |
| *Phelan-McDermid Syndrome* | 1 | 2.2 |

Twenty-five patients (55.6%) were first diagnosed and treated for catatonia in the inpatient setting by the consult-liaison psychiatry service. Twenty patients (44.4%) were recruited based on outpatient referrals. Patients were followed for a mean of 409.6 days (*SD*=256.0), with a median of 295 days [range 58–910] (Figure 1). There were 477 total outpatient appointments during the study period, of which 320 (67.1%) were conducted via telemedicine. The mean number of total appointments for each patient was 11 (*SD*=7.2), with a median of 10.

**Figure 1:**
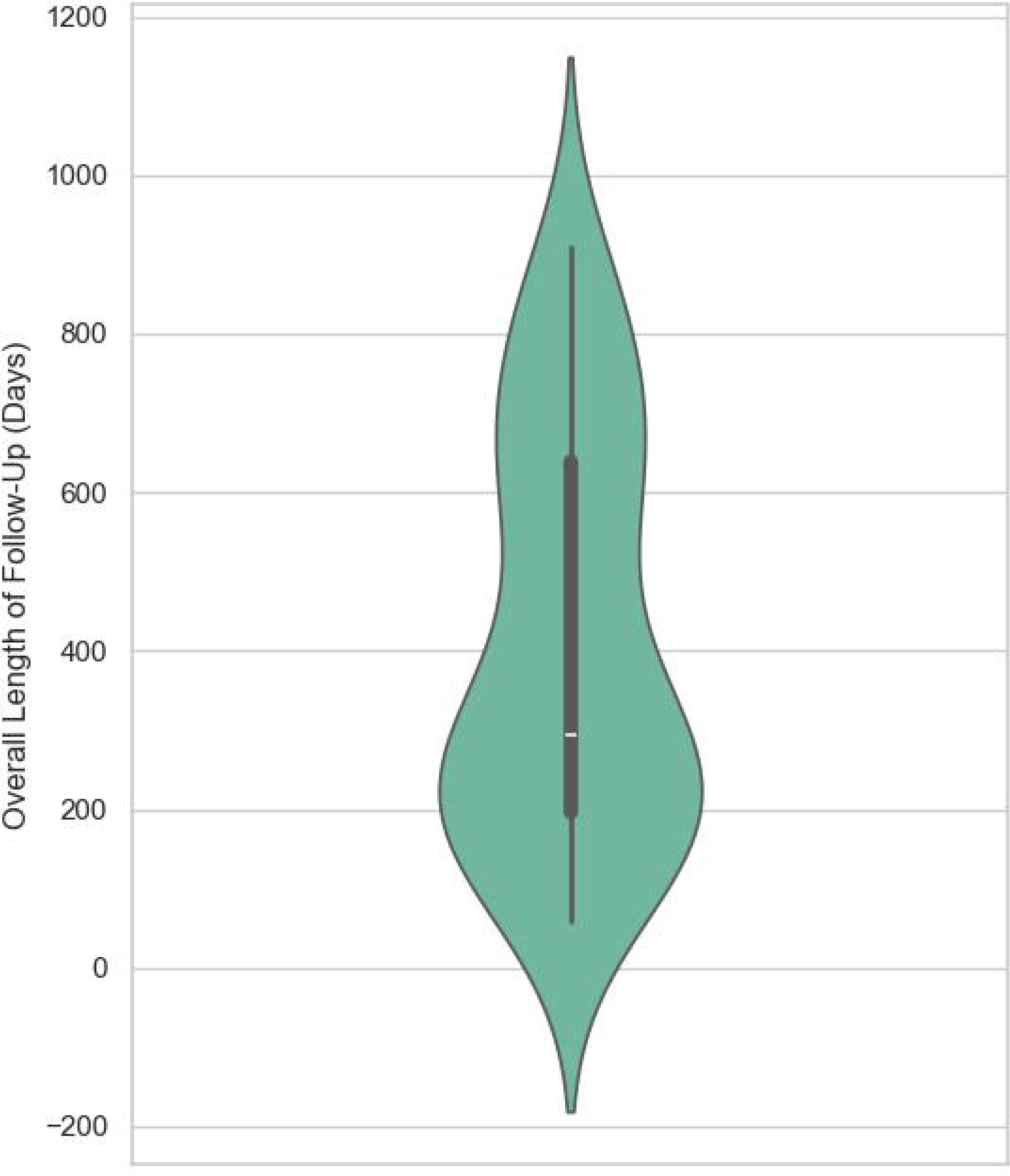
Length of Overall Outpatient Follow-Up.

### Co-occurring medical, genetic and psychiatric conditions

Co-occurring medical, genetic, and psychiatric conditions were common in this cohort (Tables 1 and 2). 20 (44.4%) had a history of seizures, 7 (15.6%) were diagnosed with epilepsy, and 2 (4.4%) patients each had prior diagnoses of delirium and Anti-N-methyl-D-aspartate (NMDA) receptor encephalitis. Additional psychiatric diagnoses were common: 35 patients (77.8%) were diagnosed with at least one psychiatric condition besides catatonia and autism. The most common of these was ADHD (*n*=25; 55.6%). Eight patients (17.8%) experienced a subjective traumatic event within one year of catatonia symptom onset.

### Frequency of index catatonic symptoms and abnormal physical examination findings

Supplemental tables 1 and 2 contain data reporting the frequency of index catatonic symptoms and physical examination findings as reported by the KCS/KCE and BFCRS, respectively. 42 (93.3%) patients had a completed index KCS/KCE and 37 (82.2%) has a completed index BFCRS. The three most common symptoms documented on the KCS included impulsivity (n=34, 80.9%), staring (n=34, 80.9%), and stereotypies (n=29, 69.0%). When explicitly measuring self-injurious mannerisms and/or stereotypies (indicated by a KCS score of 4 or more for either item), 13 patients (28.9%) presented with this symptom, with 8 (19%) patients experiencing self-injury due to stereotypies and 5 (11.9%) due to mannerisms. The three most common abnormal physical examination findings identified via the KCE were perseveration (n=24, 60%), command-verbal (n=17, 42.5%), and gibberish or verbigeration (n=14, 35%). The three most common catatonic symptom categories on the BFCRS included staring (n=29, 78.4%), impulsivity (n=28, 75.7%), and stereotypies (n=22, 58.5%).

### History of prior psychopharmacologic treatment

As per Table 2, 38 (84.4%) patients presenting for longitudinal psychopharmacologic care had a history of previous psychiatric medication trials. The mean number of failed medications was 4.8 (SD=3.6), with a median of 4 [range 1–16]. Supplemental table 3 reports medication trials by psychiatric medication class.

**Table 2:** Psychiatric co-morbidity and clinical care for patients with autism and catatonia.

|  | *N* | % |
| --- | --- | --- |
| *N* | 45 |  |
| *Psychiatric Co-morbidity* |  |  |
| *Attention-deficit hyperactivity disorder* | 25 | 55.6 |
| *Unspecified mood disorder* | 7 | 15.6 |
| *Obsessive compulsive disorder* | 7 | 15.6 |
| *Bipolar Disorder* | 4 | 8.9 |
| *Unspecified psychosis* | 2 | 4.4 |
| *Major depressive disorder* | 2 | 4.4 |
| *Impulse control disorder* | 2 | 4.4 |
| *Disruptive mood dysregulation disorder* | 2 | 4.4 |
| *Conduct disorder* | 1 | 2.2 |
| *Post-traumatic stress disorder* | 1 | 2.2 |
| *Generalized anxiety disorder* | 1 | 2.2 |
| *Subjective traumatic experience within one year* | 8 | 17.8 |
| *Total number of patients with history of psychiatric medication use* | 3 | 84.4 |
| *Number of failed psychiatric medications* |  |  |
| *Mean* |  | 4.8 (SD=3.6) |
| *Median* |  | 4 |
| *Range* |  | 1 - 16 |

### Use of alternative benzodiazepines^11,21^ in the treatment of catatonia in autism

Overall, 44 (97.8%) patients received benzodiazepines over the course of treatment. The one patient (2.2%) who did not receive benzodiazepines was treated with memantine alone. Over half of patients (*n*=24; 55.5%) were prescribed more than one benzodiazepine over the course of treatment. As seen in Table 3, over the course of the study, 33 (73.3%) patients were treated with clonazepam due to the long half-life and previously reported efficacy of clonazepam in this patient population,^11,21^ 30 (66.7%) with lorazepam, 5 (11.1%) with diazepam, and 3 (6.7%) with clobazam. 2 patients (4.4%) were prescribed both clonazepam and clobazam. In these patients, clobazam was continued due to established efficacy and historic success in controlling seizures for these patients.

**Table 3:**
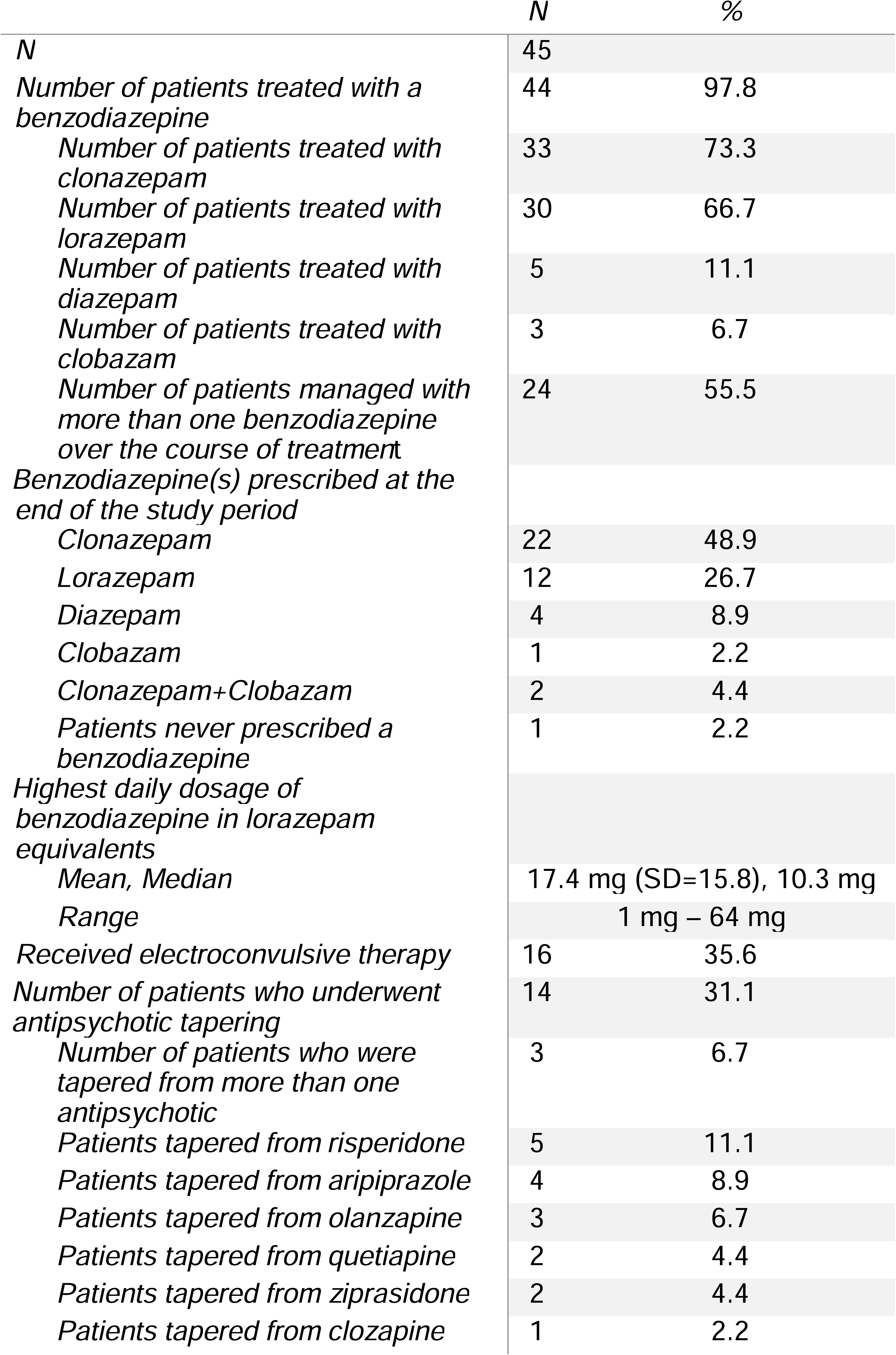
Management of benzodiazepines, electroconvulsive therapy, and antipsychotic tapering for catatonia in autism.

|  | N | % |
| --- | --- | --- |
| *N* | 45 |  |
| *Number of patients treated with a benzodiazepine* | 44 | 97.8 |
| *Number of patients treated with clonazepam* | 33 | 73.3 |
| *Number of patients treated with lorazepam* | 30 | 66.7 |
| *Number of patients treated with diazepam* | 5 | 11.1 |
| *Number of patients treated with clobazam* | 3 | 6.7 |
| *Number of patients managed with more than one benzodiazepine over the course of treatment* | 24 | 55.5 |
| *Benzodiazepine(s) prescribed at the end of the study period* |  |  |
| *Clonazepam* | 22 | 48.9 |
| *Lorazepam* | 12 | 26.7 |
| *Diazepam* | 4 | 8.9 |
| *Clobazam* | 1 | 2.2 |
| *Clonazepam+Clobazam* | 2 | 4.4 |
| *Patients never prescribed a benzodiazepine* | 1 | 2.2 |
| *Highest daily dosage of benzodiazepine in lorazepam equivalents* |  |  |
| *Mean, Median* | 17.4 mg (SD=15.8), 10.3 mg |  |
| *Range* | 1 mg – 64 mg |  |
| *Received electroconvulsive therapy* | 16 | 35.6 |
| *Number of patients who underwent antipsychotic tapering* | 14 | 31.1 |
| *Number of patients who were tapered from more than one antipsychotic* | 3 | 6.7 |
| *Patients tapered from risperidone* | 5 | 11.1 |
| *Patients tapered from aripiprazole* | 4 | 8.9 |
| *Patients tapered from olanzapine* | 3 | 6.7 |
| *Patients tapered from quetiapine* | 2 | 4.4 |
| *Patients tapered from ziprasidone* | 2 | 4.4 |
| *Patients tapered from clozapine* | 1 | 2.2 |

### Benzodiazepine dosing in lorazepam equivalents and tapering of antipsychotics and benzodiazepines

Over the course of the study period, the mean (within-individual) highest daily dose of benzodiazepines in lorazepam equivalents for the treatment of catatonia was 17.4 mg (SD=15.8) [Mdn=10.3 mg, range=1–64 mg]. Despite the high dosage of benzodiazepines, no severe adverse effects were reported, including respiratory suppression. An additional 16 (35.6%) patients were treated with ECT due to lack of response to pharmacologic treatments alone. Given the questionable efficacy of antipsychotics in catatonia (with the potential exception of clozapine),^37,38^ 14 (31.1%) patients had antipsychotic medications reduced or discontinued after the identification of catatonia. Three (6.7%) were tapered from more than one antipsychotic.

By the end of the study period, 39 patients (86.7%) remained on at least one benzodiazepine (Table 3). At the end of the study period, 22 (48.9%) were prescribed clonazepam, 12 (26.7%) lorazepam, 4 (8.9%) diazepam, 1 (2.2%) clobazam, and 2 (4.4%) were prescribed both clonazepam and clobazam. Fourteen patients (31.1%) attempted to taper off benzodiazepines during the study period; of these, 5 patients (35.7%; 11.1% of the full sample) were successfully tapered off, and the remaining 9 (64.2%; 17.8% of the full sample) discontinued the taper or resumed benzodiazepine therapy due to a return of catatonic symptoms (Table 4).

**Table 4:** Alternative pharmacologic management and benzodiazepine tapering for catatonia in autism.

|  | *N* | % |
| --- | --- | --- |
| *N* | 45 |  |
| *Number of pharmacologic agents at the end of study period* |  |  |
| *Patients on one pharmacologic agent* | 10 | 22.2 |
| *Patients on two pharmacologic agents* | 20 | 44.4 |
| *Patients on three pharmacologic agents* | 11 | 24.4 |
| *Patients on four pharmacologic agents* | 4 | 8.9 |
| *Patients on five pharmacologic agents* | 1 | 2.2 |
| *Number of patients treated with memantine* | 21 | 46.7 |
| *Mean memantine dosage* | 22.6 mg (SD=13.1) |  |
| *Number of patients treated with guanfacine* | 18 | 40.0 |
| *Mean guanfacine dosage* | 2.4 mg (SD=1.3) |  |
| *Number of patients treated with aripiprazole* | 11 | 24.4 |
| *Mean aripiprazole dosage* | 10.9 mg (SD=6.2) |  |
| *Number of patients treated with clozapine* | 6 | 13.3 |
| *Mean clozapine dosage* | 160.4 mg (SD=136.1) |  |
| *Number of patients treated with quetiapine* | 2 | 4.4 |
| *Mean quetiapine dosage* | 350 mg (SD=212.1) |  |
| *Number of patients treated with olanzapine* | 1 | 2.2 |
| *Mean olanzapine dosage* | 20 mg |  |
| *Number of patients treated with valproic acid* | 9 | 20.0 |
| *Mean valproic acid dosage* | 861.1 mg (SD=397.5) |  |
| *Number of patients treated with lithium* | 6 | 13.3 |
| *Mean lithium dosage* | 725 mg (SD=220.8) |  |
| *Number of patients who underwent a benzodiazepine taper* | 14 | 31.1 |
| *Taper discontinued due to return of catatonic symptomology* | 8 | 17.8 |
| *Number of patients successfully tapered off benzodiazepines* | 5 | 11.1 |

### Use of nonbenzodiazepine pharmacologic agents in the treatment of catatonia in autism

In addition to benzodiazepines, the use of other pharmacologic agents was common, with benzodiazepines, atypical antipsychotics, mood stabilizers, NMDA receptor antagonists, and alpha agonists used for the treatment of catatonia. Figure 2 provides a visualization of the number of psychopharmacologic medication classes used for treatment of catatonia in this cohort.

**Figure 2:**
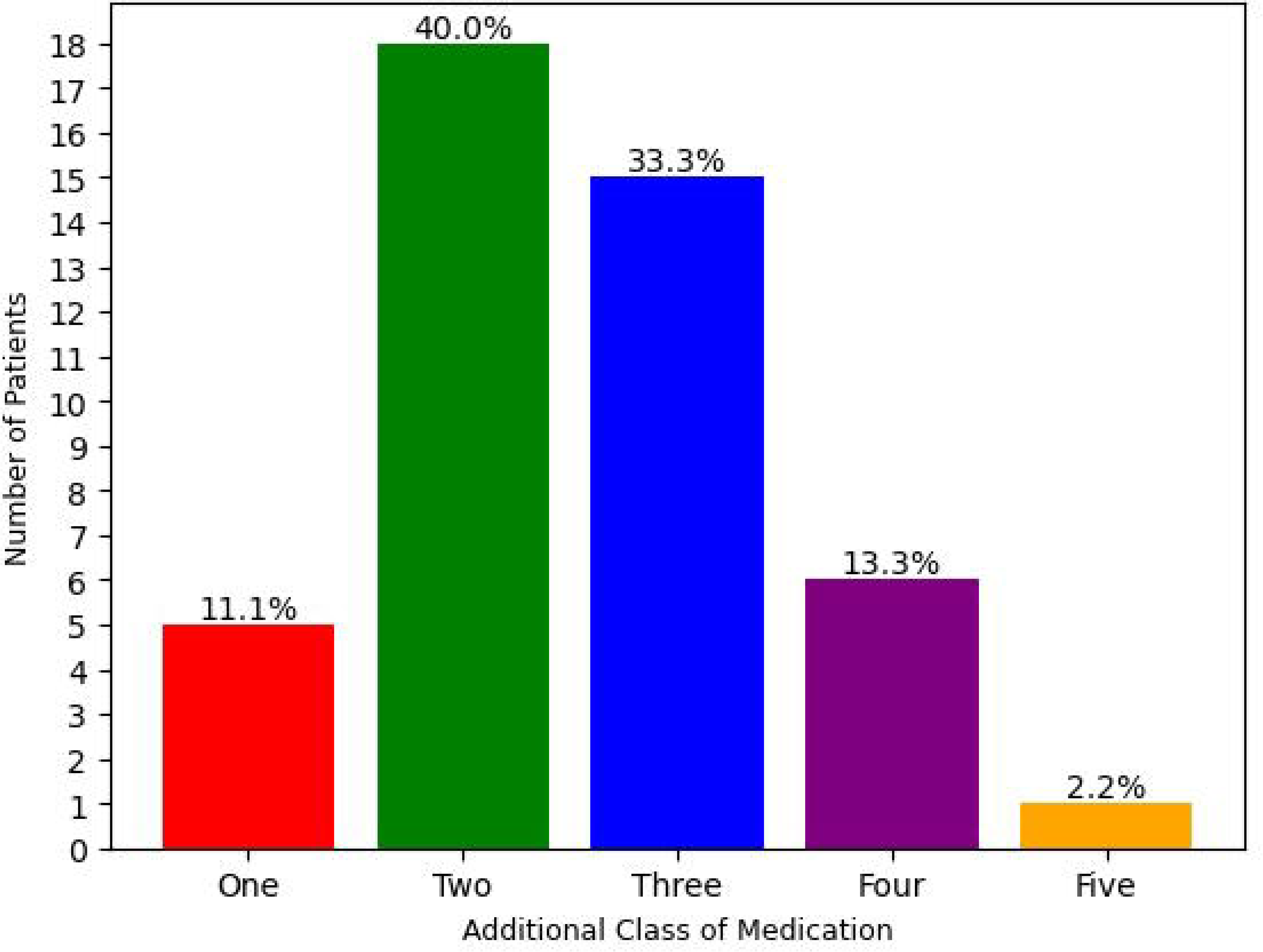
Number of Medication Classes for Catatonia in Autism Figure 3: Clinical Outcomes for Catatonia in Autism.

Of the specific nonbenzodiazepine medications used to treat catatonia in this cohort, the most frequently used were memantine (21 patients [46.7%]), guanfacine (18 patients [40%]), and aripiprazole (11 patients [24.4%]). Pharmacologic agents and doses are listed in Table 4.

### Longitudinal clinical outcomes for catatonia in autism

Of the 45 patients included in the study, 33 (73.83%) had a BFCRS, KCS, and KCE completed at the time of catatonia diagnosis or clinic intake if the patient was first diagnosed and treated for catatonia in the inpatient setting (*n*=18, 51.4%).

Over the course of the study period, the marginal mean (±SEM) BFCRS score in the sample was reduced from 13.2±1.0 to 8.7±1.0 (β=-4.5, CI_95%_ [-6.5, -2.4], *p*_adj_<.001, *d*=-0.70, CI_95%_ [-1.03, -0.38]). The marginal mean KCS reduced from 22.9±1.9 to 19.5±2.1 over the course of treatment, a difference that was not statistically significant (β=-3.4, CI_95%_ [-8.4, 1.6], *p*_adj_=.178, *d*=-0.27, CI_95%_ [-0.66, 0.12]). Lastly, the estimated marginal mean on the KCE latent trait score was reduced from –0.49±0.21 (near the estimated threshold between a “1” and “2” score on the KCE) to –1.38±0.27 (near the estimated threshold between a “0” and a “1” score on the KCE), a difference that was statistically significant *(d*_ord_=-0.88, CI_95%_ [-1.41, -0.35], *p*_adj_=.002). Lastly, the CGI-I scores *(Mdn=*2, range 1–3) were found to be significantly improved relative to a baseline (Supplemental Figure 1) score of 4 (i.e., compared to “no improvement”; *W*=0, *p*_adj_<.001, *r*_c_=-1, CI_95%_ [-1.0, -1.0]). Reductions in BFCRS, KCS, and KCE are plotted in Figure 3.

### Mortality

There were three deaths during the study period (patient ages 17, 16 and 31 years at baseline; followed for 349, 206, and 263 days, respectively). All patients were assigned male at birth and were diagnosed with profound autism. In addition, in all three patients, catatonia had improved slightly or moderately but not remitted by the time of death (catatonia-specific CGI-I ratings of 3, 2, and 2, respectively).

One patient died after choking on an excessive amount of food. This patient was receiving maintenance ECT, but their death was accidental and unrelated to ECT. Another died from recurrent pneumonia as discussed in a previous manuscript. This second patient had also received ECT, but their most recent procedure occurred three months before their death.^8^ For the final patient, the cause of death is unknown. The patient had a history of underlying cardiac pathology and developed a fever with lethargy the day before his death. The patient died the day after developing a fever. Notably, a sensitivity analysis of CGI-I scores was performed in which death (despite being ostensibly unrelated to catatonia or its treatment in these cases) was imputed as a score of 7 (“very much worse”) on the CGI-I for these three patients. In that case, the result of the CGI-I analysis was similar, albeit with a slightly smaller and less precise effect size (*W*=126, *p*_adj_<.001, *r*_c_=-.757, CI_95%_ [-1.0, -.493]).

## Discussion

To our knowledge, this study is the first observational longitudinal study to assess management and outcomes of catatonia in autism and profound autism. Patients in this cohort were followed for a median of 295 days and received extensive pharmacologic and ECT treatment.^12,14,21,22^ Despite this, most patients remained symptomatic to varying degrees over the course of the study period. Three patients died. Given the young overall age of this cohort, the high degree of morbidity and mortality provides further evidence of the substantial clinical impact of catatonia in autistic individuals.^39,40^ These findings reinforce the need for high level and specialized care for this patient population, along with high quality studies exploring further treatment options.

Similar to previous descriptive work in the field,^10^ patients in this cohort often presented with externalizing symptoms, including impulsivity, combativeness, hyperactivity, and manneristic/stereotypic self-injury. Overall, we found that over the course of clinical care there were improvements in the BFCRS, KCE, and CGI-I, all with large effect sizes. Although scores on the KCS improved in the same direction as the other measures over the course of longitudinal treatment, this change did not reach statistical significance. While numerous catatonia rating scales exist, the optimal assessment of catatonia in autistic patients remains unclear. Both the BFCRS and KCE assess for abnormal physical exam findings, while the KCS does not. The lack of improvement reported via the KCS suggests that changes in symptom burden in autistic patients with catatonia may be represented by improvement in abnormal physical examination findings associated with catatonia rather than symptom description (largely captured through an unstructured caregiver interview). Many symptoms captured in the KCS, including hyperactivity, negativism, mannerisms, stereotypies, impulsivity, incontinence, and combativeness, are present at higher rates in patients with profound autism at baseline compared to autistic individuals without profound impairment,^10^ likely due to factors other than catatonia as well. Whether specific combinations of catatonic signs or symptoms are best suited to capture improvement in catatonic features within the autistic population remains an active area of research.

The KCS, despite its creation as a developmentally sensitive measure of catatonic features, has not been rigorously psychometrically validated in the autistic population and may include a number of items that function differentially for children and adults with *profound* autism. This is especially relevant given the remarkably high prevalence of profound autism (*n*=39 [86.7%]) and autism with intellectual disability (*n*=41 [91.1%]) in this cohort. Notably, the two patients with a diagnosis of autism with intellectual disability would have *also* met criteria for profound autism if they were above 8 years of age. Consistent with previous research in pediatric catatonia, co-occurring medical and psychiatric conditions were common in our cohort.^14,41^

The prevalence of autism with intellectual disability and profound impairment in this catatonia cohort is similar to those previously reported in the literature. Specifically, previous meta-analytic work from Vaquerizo-Serrano and colleagues^10^ suggested that the rate of intellectual disability for autistic individuals with catatonia is between 5.7% and 81.6%, but reaches nearly 100% in clinic-based studies similar to the data reported in this study.^42–45^ Previous expert opinion work by Shah and colleagues has also speculated that social-emotional impairment may be the driving force behind the development of catatonia in autism.^46^ Given the high degree of correlation between social-emotional impairment and intellectual disability in autism,^47^ it is difficult to fully differentiate these co-occurring symptoms to determine which may be the primary risk factor for catatonia in autism. It has also been speculated that genetic syndromes,^48–50^ as well as traumatic or stressful experiences, may increase one’s risk for catatonia.^51,52^ Both of these factors were identified in this study, with 8 (17.8%) patients having stressful or traumatic experiences and 7 (15.5%) with co-morbid genetic syndromes. While this study does not allow us to firmly state conclusions regarding the interplay between autistic symptomology, genetic syndromes, traumatic experiences, intellectual disability, and catatonia, it suggest that there may be a cumulative effect of such factors and the overall risk of developing catatonic symptoms.^53^ Further research is urgently needed to investigate the relationship between catatonia, intellectual disability/profound impairment ^54,54,55^, and autism.

All but one patient was treated with a benzodiazepine (97.8%). Patients were most often treated initially with clonazepam or lorazepam, and over half were managed with more than one benzodiazepine. The mean highest daily dose of benzodiazepine required for treatment over the study period was 17.4 mg (SD=15.8) lorazepam equivalent; the highest daily dose recorded was 64 mg. While there is minimal literature which directly addresses benzodiazepine dosing longitudinally, this number is substantially higher than the 6 mg reported in a longitudinal hospitalization study of pediatric catatonia from our group.^19^ In autism populations specifically, doses as high as 27 mg have been reported. In these cases, symptoms of catatonia were refractory to high dose benzodiazepines but did respond to ECT.^43^

All benzodiazepines are positive allosteric modulators of the GABA-A receptor.^56^ However, differences in pharmacokinetic properties (e.g., absorption, half-life) as well as pharmacodynamic properties (GABA-A subunit specificities, potency) are present between individual agents. Thus, benzodiazepines may be expected to have varying efficacies in the treatment of catatonia. In our prior research, we found that a significant number of neurodivergent children with catatonia, who demonstrated only a partial response to lorazepam, exhibited increased responsiveness to longer-acting benzodiazepines, particularly clonazepam.^21^ One possible explanation for this finding is cortical hyperplasticity which is the hallmark of the prevailing neurobiological hypothesis of autism. It is hypothesized that an excitatory:inhibitory (E:I) imbalance is present in autism, resulting in cortical hyperactivity which may be indicative of GABAergic dysfunction and/or hyperplasticity due to impairment of long-term cortical plasticity mechanisms mediated by the NMDA receptor. ^57–59^ Recent diagnostic work in transcranial magnetic stimulation for autistic individuals without intellectual disability has reported enhanced cortical modulation, indicative of cortical hyperplasticity.^59,60^ Furthermore, recent preliminary magnetoencephalographic research has reported greater E:I imbalance in biologically male patients with autism and co-occurring intellectual disability compared to autistic individuals without intellectual disability.^61^ Overall, this suggests a direct correlation between the degree of E:I imbalance and cognitive impairment. These findings, along with the potential role of GABAergic signaling dysfunction in catatonia,^62^ may explain why longer-acting benzodiazepines at high dosages were required to stabilize catatonia for autistic individuals. Specifically, a hyperplastic cortex may rapidly acclimate to a relatively short-acting benzodiazepine such as lorazepam leading to inadequately managed catatonic symptomology.^10,59^

Tapering of benzodiazepines was attempted for 14 patients (31.1% of the sample), with nearly two thirds of those who attempted the taper (9/14 [64.2%]) resuming benzodiazepine treatment due to a return of catatonic symptoms. Thus, only 5 patients in our sample (11.1% of the full cohort) were successfully tapered off of benzodiazepines without a recurrence of catatonic symptoms. Given these challenges, further data is needed to explore the implications and possible benefits (as well as risk-benefit ratio) of long-term benzodiazepine use for patients with autism and chronic catatonia. To our knowledge, this is the first study to document attempts at benzodiazepine tapering following stabilization of catatonic symptoms.

Along with benzodiazepines, ECT has long been considered the gold-standard for treatment of catatonia across the lifecycle and spectrum of neurodivergence.^12,14,17,18^ However, access is often limited by provider availability, stigma, and state-dependent restrictive legislation which often specifically limits access for children.^16,63^ Previous research has called into question the clinical efficacy of benzodiazepines for catatonia in autism, focusing more on ECT as the primary treatment.^10,43^ However, in our study we found that the majority of patients did not require treatment with ECT (*n*=29, 64.4%) as these patients improved to the degree that ECT was no longer considered a necessity. It remains imperative to determine the utility of ECT for the treatment of medication partial-responders as data on the remission rate, recurrence risk, and long-term outcomes (including both benefits and harms) of this treatment modality remain greatly understudied.

Regarding pharmacologic interventions above and beyond the use of benzodiazepines, this study adds to the existing literature which address the use of NMDA receptor antagonists, mood stabilizers, and some antipsychotics (traditionally aripiprazole and clozapine derivatives),^21,22,38^ as well as the likely utility of using multiple pharmacologic agents in the treatment of catatonia in autism. Over 75% of the patients in this cohort required multiple medications from various pharmacologic classes for stabilization. Notably, nearly one third of the cohort was tapered from antipsychotics for the treatment of catatonia. The association of antipsychotics and catatonia is another point that is challenging to fully address. There are concerns that use of antipsychotics in the presence of catatonia may ultimately result in neuroleptic malignant syndrome (NMS), a condition associated with a high degree of morbidity and mortality.^64^ Therefore, careful consideration, as well as prior initiation of benzodiazepine treatment is warranted before initiating antipsychotics when catatonia is on the differential. However, some antipsychotics (clozapine in particular)^37^ have proven effective in managing catatonia, especially when the underlying cause of catatonia is thought to be related to a psychotic disorder,^65^ though evaluation of psychosis can be challenging for individuals with autism and intellectual disability. Overall, careful monitoring for worsening of catatonic symptoms is warranted when any antipsychotic is used in treatment.

Strengths of this study include the large sample size and longer follow-up period relative to other studies examining catatonia in autism. This study is also the first to present longitudinal outpatient pharmacologic and clinical outcome data in this population using standardized catatonia rating scales. One limitation in interpreting this data is more than half of the patients in this cohort were diagnosed with catatonia and treatment was initiated in the inpatient setting prior to entering longitudinal outpatient care. Reductions in the catatonia assessment measures are likely greatly influenced by this, as symptoms of catatonia have been shown to improve rapidly with treatment of benzodiazepines.^66,67^

The use of telemedicine in the evaluation and clinical care of these patients is also a limitation, particularly given the relevance of the physical exam to the assessment of catatonic features. At present, there are no studies which assess the sensitivity and specificity of catatonia examinations that occur over telemedicine. However, with careful involvement of family and other caregivers, telemedicine may be a viable option for catatonia assessment.^68^ Given the high frequency of aggression, self-injury, negativism, and impulsivity in this patient population, the use of telemedicine in evaluation may improve patient safety and increase inclusion of severely impacted individuals in future longitudinal catatonia clinics, and this should be studied in future prospective research. In addition, the presence of profound hyperactivity, aggression, and externalizing symptoms of catatonia may increase the difficulty in conducting a full catatonia examination. Previous research has shown that the BFCRS, KCS, and KCE can be successfully used in pediatric populations.^13,19,21^ Still, the KCE, KCS, and BFCRS are not validated in this specific patient population.^69,70^ Another limitation is the lack of specific intelligence quotient scores for patients, which could be utilized to further investigate the link between intellectual disability and catatonia. Finally, treatments were provided as part of routine clinical care and not as part of a prespecified clinical protocol, with most patients receiving multiple treatments simultaneously. As a result, this study cannot provide direct evidence for individual treatments’ effectiveness in this population.

Overall, this study provides further evidence for the substantial clinical impact of catatonia on autistic individuals, particularly those meeting the criteria for profound autism, and demonstrates the feasibility of outpatient management of catatonic symptoms. While outpatient treatment resulted in large improvements in catatonic symptoms as measured using the BFCRS, KCE, and CGI, residual symptoms remained in most patients despite receiving the gold standard of psychopharmacologic care for catatonia. Ongoing research is needed to develop improved diagnostic tools to assess catatonia in individuals with autism and to develop more effective treatments.

## Data Availability

De-identified data is available from the corresponding author (JRS) upon reasonable request

## Abbreviations

(ECT): Electroconvulsive Therapy
(STROBE): Strengthening the Reporting of Observational Studies in Epidemiology
(BFCRS): Bush Francis Catatonia Rating Scale
(KCS): Kanner Catatonia Severity
(KCE): Kanner Catatonia Examination
(BFCSI): Bush Francis Catatonia Screening Item
(CGI-I): Clinical Global Impression–Improvement

## Figure legend

**Supplemental Figure 1:**
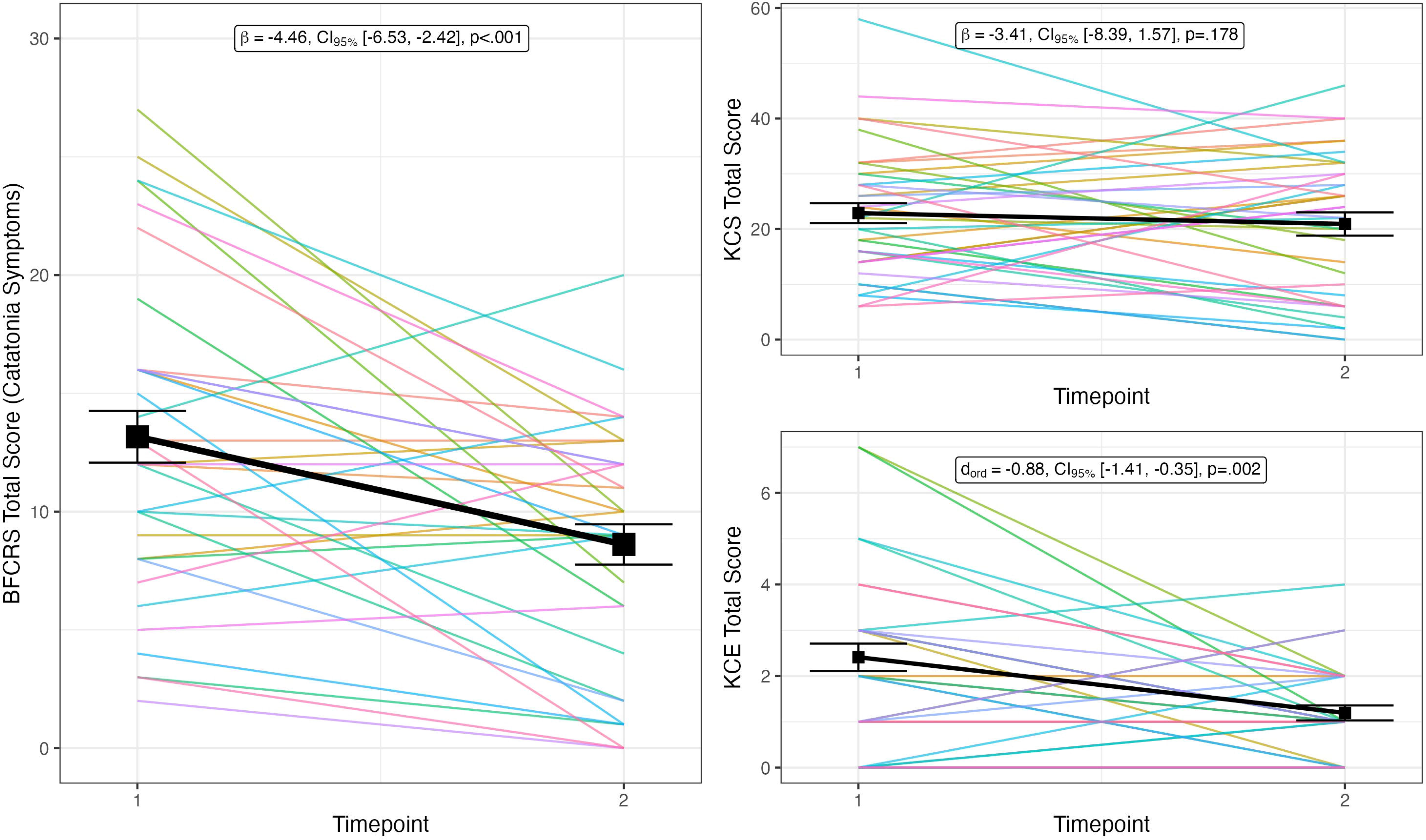
Clinical Global Impression-Improvement Scores.

## Footnotes

* We acknowledge that the term “profound autism” is controversial within the field of autism research, and its merits have been disputed by many in the autistic community, including prominent researchers and advocates.3 Furthermore, additional support-needs based classification systems have been recently proposed that may be better suited to describing this group.4 Nevertheless, in the context of the current study, we have found the descriptor of profound autism and its operational criteria to be scientifically and clinically meaningful, particularly due to the much higher likelihood of catatonia occurring in this subpopulation relative to others on the autism spectrum.

## Notes

### Competing Interest Statement

JRS receives funding from the Eunice Kennedy Shriver National Institute of Child Health and Human Development, Axial Therapeutics, and Roche. ZJW has received consulting fees from Roche and Autism Speaks. He also serves as the Vice-chair of Autistic and Neurodivergent Scholars Working for Equity in Research (ANSWER), a division of the Autism Intervention Research Network on Physical Health (AIR-P). JW receives support from the Department of Veterans Affairs, Geriatric, Research, Education and Clinical Center (GRECC) at the Tennessee Valley Healthcare System in Nashville, TN. JL receives funding from Harvard Medical School, the Rappaport Foundation, and the Foundation for Prader-Willi Research. He holds equity and has received consulting income from Revival Therapeutics, Inc.

### Funding Statement

Sources of support included grants from the Eunice Kennedy Shriver National Institute of Child Health and Human Development (1P50HD103537; JRS), the National Institute on Deafness and Other Communication Disorders (F30-DC019510; ZJW), the National Institute of General Medical Sciences (T32-GM007347; ZJW), and Vanderbilt University Medical Center Bixler-Johnson-Mayes Endowed Chair in the Department of Psychiatry and Behavioral Sciences (SL, CF). No funding agencies had any role in study design, writing of the report, or data collection, analysis, or interpretation.

### Author Declarations

This study was reviewed by the Vanderbilt University Institutional Review Board (#170317) with a waiver of informed consent from participants.

## References

1. American Psychiatric Association. *Diagnostic and Statistical Manual of Mental Disorders*. 5th, text rev ed. American Psychiatric Publishing; 2022. 10.1176/appi.books.9780890425787

2. Lord C, Charman T, Havdahl A, et al. The Lancet Commission on the future of care and clinical research in autism. The Lancet. 2022;399(10321):271–334. doi:10.1016/S0140-6736(21)01541-5

3. Kapp SK. Profound Concerns about “Profound Autism”: Dangers of Severity Scales and Functioning Labels for Support Needs. Educ Sci. 2023;13(2):106. doi:10.3390/educsci13020106

4. Sterrett K, Clarke E, Nofer J, Piven J, Lord C. Toward a functional classification for autism in adulthood. Autism Res. doi:10.1002/aur.3201

5. Koegel LK, Bryan KM, Su PL, Vaidya M, Camarata S. Definitions of Nonverbal and Minimally Verbal in Research for Autism: A Systematic Review of the Literature. J Autism Dev Disord. 2020;50(8):2957–2972. doi:10.1007/s10803-020-04402-w

6. Hughes MM, Shaw KA, DiRienzo M, et al. The Prevalence and Characteristics of Children With Profound Autism, 15 Sites, United States, 2000-2016. Public Health Rep. 2023;138(6):971–980. doi:10.1177/00333549231163551

7. Sheehan R, Mutch J, Marston L, Osborn D, Hassiotis A. Risk factors for in-patient admission among adults with intellectual disability and autism: investigation of electronic clinical records. BJPsych Open. 2020;7(1):e5. doi:10.1192/bjo.2020.135

8. Siegel M, Smith KA, Mazefsky C, et al. The autism inpatient collection: methods and preliminary sample description. Mol Autism. 2015;6:61. doi:10.1186/s13229-015-0054-8

9. Ferguson EF, Barnett ML, Goodwin JW, Vernon TW. “There is No Help:” Caregiver Perspectives on Service Needs for Adolescents and Adults with Profound Autism. J Autism Dev Disord. Published online July 4, 2024. doi:10.1007/s10803-024-06451-x

10. Vaquerizo-Serrano J, Salazar De Pablo G, Singh J, Santosh P. Catatonia in autism spectrum disorders: A systematic review and meta-analysis. Eur Psychiatry. 2022;65(1):e4. doi:10.1192/j.eurpsy.2021.2259

11. Smith JR, York T, Warn S, Borodge D, Pierce DL, Fuchs DC. Another Option for Aggression and Self-Injury, Alternative Benzodiazepines for Catatonia in Profound Autism. J Child Adolesc Psychopharmacol. 2023;33(3):109–117. doi:10.1089/cap.2022.0067

12. Rogers JP, Oldham MA, Fricchione G, et al. Evidence-based consensus guidelines for the management of catatonia: Recommendations from the British Association for Psychopharmacology. J Psychopharmacol Oxf Engl. 2023;37(4):327–369. doi:10.1177/02698811231158232

13. Smith JR, York T, Baldwin I, Fuchs C, Fricchione G, Luccarelli J. Diagnostic features of paediatric catatonia: multisite retrospective cohort study. BJPsych Open. 2024;10(3):e96. doi:10.1192/bjo.2024.61

14. Hirjak D, Rogers JP, Wolf RC, et al. Catatonia. Nat Rev Dis Primer. 2024;10(1):1–18. doi:10.1038/s41572-024-00534-w

15. Wachtel LE, Shorter E, Fink M. Electroconvulsive therapy for self-injurious behaviour in autism spectrum disorders: recognizing catatonia is key. Curr Opin Psychiatry. 2018;31(2):116–122. doi:10.1097/YCO.0000000000000393

16. Espinoza RT, Kellner CH. Electroconvulsive Therapy. N Engl J Med. 2022;386(7):667-672. doi:10.1056/NEJMra2034954

17. Smith JR, Hopkins CE, Xiong J, Luccarelli J, Shultz E, Vandekar S. Use of ECT in Autism Spectrum Disorder and/or Intellectual Disability: A Single Site Retrospective Analysis. J Autism Dev Disord. Published online December 17, 2022. doi:10.1007/s10803-022-05868-6

18. Smith JR, Baldwin I, Termini KA, McGonigle T, Vandekar S, Luccarelli J. Use of ECT for Children With and Without Catatonia: A Single-Site Retrospective Analysis. J ECT. Published online January 25, 2024. doi:10.1097/YCT.0000000000000993

19. Luccarelli J, Clauss JA, York T, et al. Exploring the Trajectory of Catatonia in Neurodiverse and Neurotypical Pediatric Hospitalizations: A Multicenter Longitudinal Analysis. medRxiv. Published online June 7, 2024:2024.06.06.24308554. doi:10.1101/2024.06.06.24308554

20. Ferrafiat V, Wachtel L, Dhossche D, Hauptman A. Catatonia is an outpatient reality. What do we do now? Schizophr Res. 2024;264:233–235. doi:10.1016/j.schres.2023.12.019

21. Smith JR, Baldwin I, York T, et al. Alternative psychopharmacologic treatments for pediatric catatonia: a retrospective analysis. Front Child Adolesc Psychiatry. 2023;2:1208926. doi:10.3389/frcha.2023.1208926

22. Beach SR, Gomez-Bernal F, Huffman JC, Fricchione GL. Alternative treatment strategies for catatonia: A systematic review. Gen Hosp Psychiatry. 2017;48:1–19. doi:10.1016/j.genhosppsych.2017.06.011

23. von Elm E, Altman DG, Egger M, et al. The Strengthening the Reporting of Observational Studies in Epidemiology (STROBE) statement: guidelines for reporting observational studies. J Clin Epidemiol. 2008;61(4):344–349. doi:10.1016/j.jclinepi.2007.11.008

24. Louie ATH, Anand E, Baldwin I, Smith JR. Use of En-Bloc Multiple Monitored Electroconvulsive Therapy in Benzodiazepine Refractory Malignant Catatonia. J ECT. Published online June 28, 2024. doi:10.1097/YCT.0000000000001020

25. Bush G, Fink M, Petrides G, Dowling F, Francis A. Catatonia. I. Rating scale and standardized examination. Acta Psychiatr Scand. 1996;93(2):129-136. doi:10.1111/j.1600-0447.1996.tb09814.x

26. Carroll BT, Kirkhart R, Ahuja N, et al. Katatonia. Psychiatry Edgmont. 2008;5(12):42–50.

27. Bush G, Fink M, Petrides G, Dowling F, Francis A. Catatonia. I. Rating scale and standardized examination. Acta Psychiatr Scand. 1996;93(2):129-136. 10.1111/j.1600-0447.1996.tb09814.x

28. Weleff J, Barnett BS, Park DY, Akiki TJ, Aftab A. The State of the Catatonia Literature: Employing Bibliometric Analysis of Articles From 1965-2020 to Identify Current Research Gaps. J Acad Consult-Liaison Psychiatry. 2023;64(1):13–27. doi:10.1016/j.jaclp.2022.07.002

29. Busner J, Targum SD. The Clinical Global Impressions Scale. Psychiatry Edgmont. 2007;4(7):28–37.

30. Benzodiazepine Conversion Calculator. MDCalc. Accessed August 20, 2024. https://www.mdcalc.com/calc/10091/benzodiazepine-conversion-calculator

31. Benzodiazepine Equivalents Conversion Calculator - ClinCalc.com. Accessed August 20, 2024. https://clincalc.com/Benzodiazepine/

32. Weleff J, Barnett BS, Park DY, Akiki TJ, Aftab A. The State of the Catatonia Literature: Employing Bibliometric Analysis of Articles From 1965-2020 to Identify Current Research Gaps. J Acad Consult-Liaison Psychiatry. 2023;64(1):13–27. doi:10.1016/j.jaclp.2022.07.002

33. Hirjak D, Brandt GA, Fritze S, Kubera KM, Northoff G, Wolf RC. Distribution and frequency of clinical criteria and rating scales for diagnosis and assessment of catatonia in different study types. Schizophr Res. 2024;263:93–98. doi:10.1016/j.schres.2022.12.019

34. Bates D, Mächler M, Bolker B, Walker S. Fitting Linear Mixed-Effects Models Using lme4. J Stat Softw. 2015;67:1–48. doi:10.18637/jss.v067.i01

35. Bürkner PC, Vuorre M. Ordinal Regression Models in Psychology: A Tutorial. Adv Methods Pract Psychol Sci. 2019;2(1):77–101. doi:10.1177/2515245918823199

36. Christensen RHB. ordinal: Regression Models for Ordinal Data. Published online August 19, 2024. Accessed August 20, 2024. https://cran.r-project.org/web/packages/ordinal/index.html

37. Saini A, Begum N, Matti J, et al. Clozapine as a treatment for catatonia: A systematic review. Schizophr Res. 2024;263:275–281. doi:10.1016/j.schres.2022.09.021

38. Thom RP, Wu M, Ravichandran C, McDougle CJ. Clozapine for treatment refractory catatonia in individuals with autism spectrum disorder: a retrospective chart review study. Expert Rev Clin Pharmacol. 2023;16(9):865–875. doi:10.1080/17512433.2023.2243820

39. Cornic F, Consoli A, Tanguy ML, et al. Association of adolescent catatonia with increased mortality and morbidity: evidence from a prospective follow-up study. Schizophr Res. 2009;113(2-3):233–240. doi:10.1016/j.schres.2009.04.021

40. Hirvikoski T, Mittendorfer-Rutz E, Boman M, Larsson H, Lichtenstein P, Bölte S. Premature mortality in autism spectrum disorder. Br J Psychiatry J Ment Sci. 2016;208(3):232–238. doi:10.1192/bjp.bp.114.160192

41. Luccarelli J, Kalinich M, Fricchione G, Smith F, Beach SR, Smith JR. Diagnostic and demographic factors of pediatric and adult catatonia hospitalizations: A 2016-2020 National Inpatient Sample Study. Acta Psychiatr Scand. Published online August 8, 2024. doi:10.1111/acps.13744

42. Wing L, Shah A. Catatonia in autistic spectrum disorders. Br J Psychiatry J Ment Sci. 2000;176:357–362. doi:10.1192/bjp.176.4.357

43. Wachtel LE. Treatment of catatonia in autism spectrum disorders. Acta Psychiatr Scand. 2019;139(1):46–55. doi:10.1111/acps.12980

44. Wing L, Shah A. A Systematic Examination of Catatonia-like Clinical Pictures in Autism Spectrum Disorders. In: Dhossche DM, Wing L, Ohta M, Neumärker K, eds. International Review of Neurobiology. Vol 72. Catatonia in Autism Spectrum Disorders. Academic Press; 2006:21–39. doi:10.1016/S0074-7742(05)72002-X

45. Ohta M, Kano Y, Nagai Y. Catatonia in individuals with autism spectrum disorders in adolescence and early adulthood: a long-term prospective study. Int Rev Neurobiol. 2006;72:41–54. doi:10.1016/S0074-7742(05)72003-1

46. Shah A. *Catatonia, Shutdown and Breakdown in Autism: A Psycho-Ecological Approach*. Jessica Kingsley Publishers; 2019.

47. Bernard Paulais MA, Mazetto C, Thiébaut E, et al. Heterogeneities in Cognitive and Socio-Emotional Development in Children With Autism Spectrum Disorder and Severe Intellectual Disability as a Comorbidity. Front Psychiatry. 2019;10:508. doi:10.3389/fpsyt.2019.00508

48. Raffin M, Consoli A, Giannitelli M, et al. Catatonia in Children and Adolescents: A High Rate of Genetic Conditions. J Am Acad Child Adolesc Psychiatry. 2018;57(7):518–525.e1. doi:10.1016/j.jaac.2018.03.020

49. Moore S, Amatya DN, Chu MM, Besterman AD. Catatonia in autism and other neurodevelopmental disabilities: a state-of-the-art review. Npj Ment Health Res. 2022;1(1):12. doi:10.1038/s44184-022-00012-9

50. Shillington A, Zappia KJ, White L, et al. Genetic syndromes are prevalent in patients with comorbid neurodevelopmental disorders and catatonia. Am J Med Genet A. 2023;191(11):2716–2722. doi:10.1002/ajmg.a.63379

51. Fink M, Taylor MA. *Catatonia: A Clinician’s Guide to Diagnosis and Treatment*. Cambridge University Press; 2006.

52. Dhossche DM, Ross CA, Stoppelbein L. The role of deprivation, abuse, and trauma in pediatric catatonia without a clear medical cause. Acta Psychiatr Scand. 2012;125(1):25–32. doi:10.1111/j.1600-0447.2011.01779.x

53. Waizbard-Bartov E, Fein D, Lord C, Amaral DG. Autism severity and its relationship to disability. Autism Res. 2023;16(4):685–696. doi:10.1002/aur.2898

54. Stedman A, Taylor B, Erard M, Peura C, Siegel M. Are Children Severely Affected by Autism Spectrum Disorder Underrepresented in Treatment Studies? An Analysis of the Literature. J Autism Dev Disord. 2019;49(4):1378–1390. doi:10.1007/s10803-018-3844-y

55. Russell PSS. The clinical utility of a multivariate genetic panel for identifying those at risk of developing Opioid Use Disorder while on prescription opioids. Scand J Pain. Published online December 27, 2019. doi:10.1515/sjpain-2019-0160

56. Griffin CE, Kaye AM, Bueno FR, Kaye AD. Benzodiazepine Pharmacology and Central Nervous System–Mediated Effects. Ochsner J. 2013;13(2):214–223.

57. Casanova MF, Shaban M, Ghazal M, et al. Effects of Transcranial Magnetic Stimulation Therapy on Evoked and Induced Gamma Oscillations in Children with Autism Spectrum Disorder. Brain Sci. 2020;10(7):423. doi:10.3390/brainsci10070423

58. Rojas DC. The role of glutamate and its receptors in autism and the use of glutamate receptor antagonists in treatment. J Neural Transm. 2014;121(8):891–905. doi:10.1007/s00702-014-1216-0

59. Smith JR, DiSalvo M, Green A, et al. Treatment Response of Transcranial Magnetic Stimulation in Intellectually Capable Youth and Young Adults with Autism Spectrum Disorder: A Systematic Review and Meta-Analysis. Neuropsychol Rev. Published online September 26, 2022. doi:10.1007/s11065-022-09564-1

60. Jannati A, Ryan MA, Kaye HL, Tsuboyama M, Rotenberg A. Biomarkers Obtained by Transcranial Magnetic Stimulation in Neurodevelopmental Disorders. J Clin Neurophysiol. 2022;39(2):135–148. doi:10.1097/WNP.0000000000000784

61. Manyukhina VO, Prokofyev AO, Galuta IA, et al. Globally elevated excitation-inhibition ratio in children with autism spectrum disorder and below-average intelligence. Mol Autism. 2022;13(1):20. doi:10.1186/s13229-022-00498-2

62. Walther S, Stegmayer K, Wilson JE, Heckers S. Structure and neural mechanisms of catatonia. Lancet Psychiatry. 2019;6(7):610–619. doi:10.1016/S2215-0366(18)30474-7

63. Ong M, Patterson E, Stewart L, Pierce D, Smith JR. Morbidity Due to Disparity in Pediatric Electroconvulsive Therapy. J Am Acad Child Adolesc Psychiatry. 2023;62(3):279–281. doi:10.1016/j.jaac.2022.07.850

64. Connell J, Oldham M, Pandharipande P, et al. Malignant Catatonia: A Review for the Intensivist. J Intensive Care Med. 2023;38(2):137–150. doi:10.1177/08850666221114303

65. Patterson EM, Lim J, Fuchs P, Smith JR, Moussa-Tooks A, Ward HB. Use of First-Generation Antipsychotics in an Adolescent Male with Catatonic Schizophrenia. Harv Rev Psychiatry. Published online October 13, 2023. doi:10.1097/HRP.0000000000000381

66. Luccarelli J, McCoy TH, York T, et al. The effectiveness of the lorazepam challenge test in pediatric catatonia: A multisite retrospective cohort study. Schizophr Res. 2024;270:410–415. doi:10.1016/j.schres.2024.07.004

67. Suchandra HH, Reddi VSK, Aandi Subramaniyam B, Muliyala KP. Revisiting lorazepam challenge test: Clinical response with dose variations and utility for catatonia in a psychiatric emergency setting. Aust N Z J Psychiatry. 2021;55(10):993–1004. doi:10.1177/0004867420968915

68. Luccarelli J, Fricchione G, Newton AW, Wozniak J. The diagnosis and treatment of catatonia via telemedicine: A case report and proposed diagnostic criteria. Schizophr Res. 2022;241:66–67. doi:10.1016/j.schres.2022.01.038

69. Erdoğan İM, Aytulun A, Avanoğlu KB, et al. Turkish Adaptation, Validity and Reliability Study of the Bush Francis Catatonia Rating and KANNER Scales. Turk J Psychiatry. 2022;34(4):254–261. doi:10.5080/u27064

70. Wilson JE, Niu K, Nicolson SE, Levine SZ, Heckers S. The diagnostic criteria and structure of catatonia. Schizophr Res. 2015;164(1-3):256–262. doi:10.1016/j.schres.2014.12.036

