## Supplemental Tables for "Longitudinal Symptom Burden and Pharmacologic Management of Catatonia in Autism with and without Profound Impairment: An Observational Study"

**Supplemental Table 1:** Frequency of index catatonic symptoms in autism as measured by the Kanner Catatonia Severity Scale and Kanner Catatonia Examination

|  | N | % |
| --- | --- | --- |
| N | 42 |  |
| Kanner Catatonia Severity Scale |  |  |
| Excitement-hyperactivity | 26 | 61.9 |
| Immobility | 12 | 28.5 |
| Stupor | 4 | 9.5 |
| Mutism | 18 | 42.9 |
| Staring | 34 | 80.9 |
| Posturing | 13 | 30.9 |
| Grimacing | 23 | 54.8 |
| Stereotypy | 29 | 69.0 |
| Stereotypy with self-injury (score of four or higher) | 8 | 19.0 |
| Mannerisms | 21 | 50.0 |
| Mannerisms with self-injury (score of four or higher) | 5 | 11.9 |
| Rigidity | 18 | 42.9 |
| Flaccidity | 0 | 0 |
| Negativism | 28 | 66.7 |
| Reduced food intake | 5 | 11.9 |
| Reduced fluid intake | 5 | 11.9 |
| Impulsivity | 34 | 80.9 |
| Nudism | 3 | 7.1 |
| Incontinence | 18 | 42.9 |
| Combativeness | 24 | 57.1 |
| Kanner Catatonia Examination | 40 |  |
| Parroting (Echolalia) | 8 | 20.0 |
| Gibberish (Verbigeration) | 14 | 35.0 |
| Perseveration | 24 | 60.0 |
| Waxy flexibility | 2 | 5.0 |
| Catalepsy | 3 | 7.5 |
| Echopraxia | 6 | 15.0 |
| Command-verbal (Automatic obedience and ambitendency) | 17 | 42.5 |
| Command-motor (Mitgehen) | 5 | 12.5 |
| Paratonia (Gegenhalten) | 3 | 7.5 |
| Grasp reflex | 2 | 5.0 |
| Metronome test | 2 | 5.0 |
| Magnetism | 8 | 20.0 |

**Supplemental Table 2**: Frequency of index catatonic symptoms in autism as measured by the Bush Francis Catatonia Rating Scale

|  | N | % |
| --- | --- | --- |
| N | 37 |  |
| Excitement | 21 | 56.8 |
| Immobility | 10 | 27.0 |
| Mutism | 14 | 37.8 |
| Staring | 29 | 78.4 |
| Posturing | 10 | 27.0 |
| Grimacing | 18 | 48.6 |
| Echolalia/echopraxia | 13 | 35.1 |
| Stereotypy | 22 | 58.5 |
| Mannerisms | 16 | 43.2 |
| Verbigeration | 10 | 27.0 |
| Rigidity | 17 | 45.9 |
| Negativism | 20 | 54.1 |
| Waxy flexibility | 1 | 2.7 |
| Withdrawal | 6 | 16.2 |
| Impulsivity | 28 | 75.7 |
| Automatic obedience | 8 | 21.6 |
| Mitgehen | 5 | 13.5 |
| Gegenhalten | 1 | 2.7 |
| Ambitendency | 11 | 29.7 |
| Grasp reflex | 3 | 8.1 |
| Perseveration | 16 | 43.2 |
| Combativeness | 14 | 37.8 |
| Autonomic instability | 10 | 27.0 |

**Supplemental Table 3**: Number of failed psychiatric medications per class

|  |  | N | % | | |
| --- | --- | --- | --- | --- | --- |
|  | N | 45 |  | | |
|  | Antidepressants | 24 | 53.3 | | |
|  | Mean number of failed trials | 1.6 (SD=0.9) | | | |
|  | Second generation antipsychotics | 21 | 46.7 | | |
|  | Mean number of failed trials | 2.2 (SD=1.1) | | | |
|  | Alpha agonists | 19 | 42.2 | | |
|  | Mean number of failed trials | 1.1 (SD=0.3) | | | |
|  | Stimulants | 14 | | | 31.1 |
|  | Mean number of failed trials | 1.4 (SD=0.7) | | | |
|  | Mood stabilizers | 6 | | 13.3 | |
|  | Mean number of failed trials | 1.3 (SD=0.5) | | | |
|  | First generation antipsychotics | 5 | | 11.1 | |
|  | Mean number of failed trials | 1.2 (SD=0.8) | | | |
|  | NMDA receptor antagonists | 4 | | 8.9 | |
|  | Mean number of failed trials | 1 | | | |
|  | Antihistamine medication | 4 | | 8.9 | |
|  | Mean number of failed trials | 1 | | | |
| Beta-blocker medication | | 2 | | 4.4 | |
| Mean number of failed trials | | 1 | | | |
