## Supplementary figures and images for "Longitudinal Symptom Burden and Pharmacologic Management of Catatonia in Autism with and without Profound Impairment: An Observational Study"

### Supplemental Figure 1

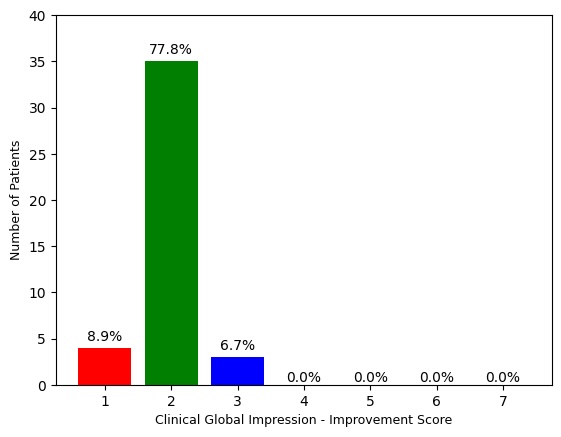
